# Automating ACMG Variant Classifications With BIAS-2015 v2.0.0: Algorithm Analysis and Benchmark Against the FDA-Approved eRepo Dataset

**DOI:** 10.1101/2025.02.23.25322737

**Authors:** Chris Eisenhart, Rachel Brickey, Brian Nadon, Joel Mewton, Vafa Bayat

## Abstract

**Background:** In 2015, the American College of Medical Genetics and Genomics (ACMG), in collaboration with the Association of Molecular Pathologists (AMP), published guidelines for interpreting and classifying germline genomic variants. These guidelines defined five categories: benign, likely benign, uncertain significance, likely pathogenic, and pathogenic, with 28 criteria but no specific implementation algorithms.

**Results:** Here we present Bitscopic Interpreting ACMG Standards 2015 (BIAS-2015 v2.0.0), an open-source software that automates the classification of variants based on 19 ACMG criteria while enabling user-defined weighting and manual adjustments for clinical contexts. BIAS-2015 supports high-throughput classification via command line, along with a web-based GUI, enabling variant review, modification, and interactive curation. Using genomic data from the FDA-recognized ClinGen Evidence Repository (eRepo v2.2.0), we evaluated BIAS-2015’s sensitivity, specificity, and F1 values with expert curation. BIAS-2015 demonstrated superior performance to InterVar, achieving a pathogenic sensitivity of 73.99% (vs. 64.31%), benign sensitivity of 80.23% (vs. 53.91%), and a 11x speed improvement, classifying 1,327 variants per second.

**Conclusion:** BIAS-2015 provides an accurate, scalable, and transparent ACMG classification framework. All code and the interactive variant curation platform are available on GitHub.

## Introduction

The clinical interpretation of genetic variants is crucial for translating genomic data into actionable clinical practice. In 2015, the American College of Medical Genetics and Genomics (ACMG) and the Association for Molecular Pathologists (AMP) established comprehensive guidelines for the interpretation and classification of germline genomic variants into five categories: pathogenic (P), likely pathogenic (LP), uncertain significance (VUS), likely benign (LB), and benign (B) [28]. These guidelines are based on 28 criteria grouped into different levels of evidence, such as population data, computational predictions, functional data, segregation data, and *de novo* data. Over the years, these guidelines have become the community standard due to their systematic approach [19].

Despite these guidelines, the interpretation of genetic variants remains a complex and labor-intensive process [18, 29]. Since 2015, many tools have been created to automate this process, including InterVar, Sherloc, PathoMAN, and others [27, 12, 33]. When deciding to create BIAS-2015, we evaluated several factors, including the need for open-source code bases, algorithms that function on freely accessible data and software, adherence to ACMG classifiers and combining criteria, ability to up and down weight all codes, and tools that report detailed explanations for each classifier code. In our analysis of available software, we did not find any that met all our criteria. Thus, we set out to create BIAS-2015.

BIAS-2015 is built on top of Illumina’s Nirvana 3.18.1 software, which is freely available under a GNU GPL license on GitHub [21]. Nirvana is a command-line annotation tool that processes VCF files into JSON format, serving as the input for BIAS-2015. Unlike other classification tools that impose licensing fees, restrictive registrations, or require proprietary infrastructure, BIAS-2015 remains fully accessible. It can be downloaded and run on any standard VCF file, on any standard computer setup, without specialized hardware or locked-down dependencies. BIAS-2015 supports input files annotated with either hg19 or hg38 genome builds, with our analysis conducted using hg19.

The algorithm operates in three phases: the pre-processing step, which gathers data from community-standard sources and extracts the information needed for classification; the data loading step, which loads datasets generated during pre-processing; and the classification phase, where ACMG codes are applied, and the combining criteria are used to classify each variant.

BIAS-2015 also supports user-provided classifiers through an optional external call file. This feature enables users to input values for the 9 ACMG classifiers that BIAS-2015 does not evaluate, adjust weights for each classifier, or overwrite BIAS-2015-generated classifier values.

To enhance flexibility and autonomy, the pre-processing phase of BIAS-2015 is included as part of the open-source software, allowing users to update datasets independently. By decoupling pre-processing from the core pipeline, users can incorporate new community-standard datasets or update existing ones as they become available. This design ensures that researchers and clinicians can reanalyze their data on their own timeline without waiting for external updates or intervention from the BIAS-2015 development team. This capability is particularly critical in a rapidly evolving field, where updated evidence or databases can significantly impact variant interpretation [22].

BIAS-2015 utilizes the ClinGen eRepo dataset as a primary reference for benchmarking classification performance [3]. To integrate eRepo into our pipeline, we downloaded the dataset and converted it into a standardized VCF format using a custom script, which is available in our GitHub repository. This conversion was performed using GenBank and RefSeq accessions from the NCBI genome database [6] and the hg19 human genome reference to ensure compatibility with downstream analyses. The resulting VCF file was then used as an input dataset for comparison and validation of BIAS-2015’s classification accuracy.

Finally, we directly compared Nirvana & BIAS-2015 to Annovar & InterVar, another ACMG classification pipeline, using the same ClinGen eRepo dataset [34, 27]. BIAS-2015 outperformed InterVar across nearly all key metrics, achieving a pathogenic sensitivity of 73.99% (vs. 64.31%), pathogenic specificity of 88.07% (vs. 90.06%), benign sensitivity of 80.23% (vs. 53.91%), and benign specificity of 96.56% (vs. 96.52%). Additionally, BIAS-2015 processed 1,327 variants per second, an 11x improvement over InterVar’s 120 variants per second.

To further support real-world variant curation, BIAS-2015 includes an interactive web application for manual review and classification adjustments. This GUI enables researchers and clinicians to refine classifications, document rationale, and efficiently manage variant interpretation workflows. By integrating a scalable classification algorithm with an interactive user interface, BIAS-2015 provides a comprehensive platform for automated and manual ACMG variant interpretation.

## Implementation

### Algorithm Overview

BIAS-2015 can be run beginning with any standard variant call format (VCF) file. First the file is annotated with Nirvana, then the resulting JSON file is passed into BIAS-2015 v2.0.0. BIAS-2015 v2.0.0 outputs a TSV file with one row per variant, providing the ACMG classification and detailed rationales for every code that was applied.

The BIAS-2015 algorithm operates in three distinct phases, ensuring both accuracy and efficiency in variant classification (Figure 1):

**Fig. 1.**
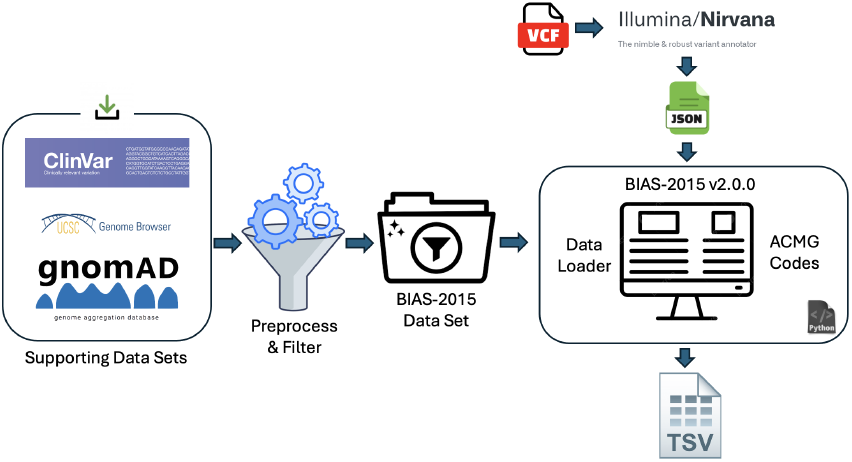
BIAS-2015 v2.0.0 Workflow. The pipeline integrates multiple genomic data sources, including ClinVar, gnomAD, and UCSC Genome Browser, which are preprocessed and filtered into the BIAS-2015 dataset. Variant annotation is performed using Illumina Nirvana, generating structured JSON files. The BIAS-2015 v2.0.0 system then processes these datasets, applies ACMG classification criteria, and outputs a final classification TSV file.

#### 1. Preprocessing Phase

BIAS-2015 gathers and formats data from community-standard sources, such as the UCSC Genome Browser FTP site [35]. This computationally intensive step extracts and optimizes relevant information for rapid runtime classification, requiring significantly more time than the classification phase.

#### 2. Data Loading Phase

At the start of runtime BIAS-2015 loads the preprocessed datasets into memory using optimized Python data structures. This eliminates later needs for runtime disk access and ensures all necessary information is readily available, enabling high-speed classification.

#### 3. Classification Phase

BIAS-2015 sequentially processes variants from the JSON file, applying 19 ACMG criteria (e.g., PVS1, PS1, PM2, BP4) using pre-loaded data [28]. The ACMG combining criteria are then used to assign a final classification, achieving high throughput with minimal computational overhead.

### Preprocessing and Data File Generation

The ACMG standards and the broader clinical bioinformatics community emphasize the importance of tools that can be updated and re-run as new genomic discoveries are made [22]. To address this need, the BIAS-2015 preprocessing process is fully automated and reproducible, allowing users to generate or update the data files required for classification. This ensures compatibility with evolving datasets and enables users to integrate evidence from their own projects or laboratories.

The preprocessing.py script automates data preparation, downloading, and formatting for use in BIAS-2015. It supports both hg19 and hg38 genome builds and simplifies the otherwise time-intensive preprocessing step. The script runs with a single command, taking approximately two hours, downloading 20 GB of data, and requiring up to 8 GB of memory. Key datasets include NCBI RefSeq HGMD Data for PVS1 Caveat, ABSplice Data for PVS1, PP3, BP4, and BP7, ClinVar VCF Data for PS1 [23], AVADA Big Bed Data for PS3, GWAS Catalog for PS4 [17], Pathogenic Domains for PM1, RepeatMasker and CCDS Data for PM4 and BP1 [13, 1], and Missense Pathogenic and Truncating Genes for PP2 and BP1.

These datasets are tailored for 11 ACMG classifiers; PVS1, PP3, BP4, BP7, PS1, PS3, PS4, PM1, PM4, BP1, and PP2. Users can customize the pre-processing step to include additional evidence by modifying source files, such as adding lab specific variants, or including new genes. This flexibility allows seamless incorporation of new data, or proprietary data, without requiring updates from the BIAS-2015 development team.

### Nirvana Data Extraction

BIAS-2015 leverages the Nirvana annotations throughout the classification process [21]. This is done by extracting the necessary information from the Nirvana JSON into a Python data structure that represents a variant, then using this information during classification. BIAS-2015 extracts and uses the following information for each variant: chromosome, position, reference allele, alternate allele, variant type, amino acid change, gene name, consequence, top ClinVar entry, PubMed ID list, ClinGen gene associated disease, dbSNP ID list, canonical transcript, full RefSeq transcript list, gnomAD, TopMed, 1000 Genomes Project, phyloP, GERP, DANN and REVEL scores [4, 8, 4, 25, 14, 23, 7, 30, 16, 15, 26]. Please see the Nirvana documentation for a detailed explanation of how they define and apply these annotations to each variant.

### Classifier Scoring and Weights

The ACMG guidelines define a framework for variant classification using multiple lines of evidence, ranging from population data to computational predictions and functional studies [28]. These criteria are assigned qualitative strength levels such as supporting, moderate, and very strong. However, the strength of individual criteria can vary depending on gene and variant specific factors, requiring careful weighting to ensure accurate classification. While many variant classification tools apply fixed weights based on ACMG recommended defaults, evolving best practices suggest that criteria should be adjusted dynamically based on the strength of supporting evidence [9].

BIAS-2015 implements a novel approach by weighting ACMG criteria dynamically at runtime, rather than relying solely on static assignments. Each criterion is assigned to one of five levels—stand-alone, very strong, strong, moderate, or supporting—based on the level of evidence supporting the call. This is achieved by assigning an integer score (1-5) to each classifier, where 1 corresponds to supporting and 5 to stand-alone. For example, PVS1 (predicted loss-of-function) is typically weighted as very strong (score of 4), but in BIAS-2015, it is down-weighted to moderate (score of 3) in genes where LOEUF of greater than 1, as these genes are tolerant to deletion and a very strong classification is not applicable [14]. Conversely, PP3 (computational evidence supporting pathogenicity) can be up-weighted to moderate or strong if multiple independent in silico tools provide consistent, high-confidence pathogenic predictions [32]. This approach ensures that variant classification reflects the most accurate and contextually relevant weighting while remaining aligned with best-practice variant classification established by expert panels [3].

Additionally, BIAS-2015 supports user defined modifications, allowing manual adjustments to classifier weights based on expert review, lab specific practices, or specific clinical considerations. Users can override the automatically assigned ACMG classifications through a structured external call file, enabling precise adjustments for cases where additional evidence or expert judgment is required. Furthermore, the BIAS-2015 web based UI provides an interactive interface for manual classification review and real time modifications. This interface allows users to inspect variant classifications, modify supporting criteria weights, and document rationales for adjustments, ensuring full transparency in variant interpretation. These features allow BIAS-2015 to function both as a high throughput automated classifier and as a flexible platform for expert guided curation.

### ClinVar Review Evidence Weights

BIAS-2015 extensively utilizes ClinVar data during the preprocessing phase to generate classification reference files [23]. To ensure robust classification, BIAS-2015 applies weighted scoring to ClinVar variants based on their review status. Variants with the highest confidence—those marked as ‘Practice Guidelines’ or ‘Reviewed by Expert Panel’—are assigned the strongest weights, whereas variants with a review status of ‘Criteria provided, conflicting interpretations’ receive reduced weight. Variants lacking assertion criteria are assigned a weight of zero and are excluded from classification to prevent unreliable data from influencing variant interpretation.

All ClinVar-derived data files used by BIAS-2015 contain explicit review-based weighting, which is directly integrated into the classification process. These weights are mapped to the standardized Sequence Variant Interpretation (SVI) framework for ACMG classification, aligning with the stand-alone, very strong, strong, moderate, and supporting evidence categories. A supplementary file provides a detailed breakdown of the ClinVar review statuses and their corresponding weights in BIAS-2015. This approach ensures a consistent, evidence-based evaluation of ClinVar annotations across all classifications.

### LOEUF Adjusted Population Cutoffs

BIAS-2015 integrates LOEUF (Loss-of-Function Observed/Expected Upper Bound Fraction) scores to refine allele frequency thresholds based on gene constraint [14]. Genes with lower LOEUF scores are highly constrained, meaning pathogenic variants in these genes tend to be rare. Conversely, genes with higher LOEUF scores are more tolerant to variation, allowing for higher pathogenic allele frequencies.

To reflect this, BIAS-2015 assigns allele frequency cutoffs based on five LOEUF-defined bins, adjusting pathogenicity thresholds accordingly (see Supplemental Data). More constrained genes receive stricter frequency cutoffs, while more tolerant genes receive more lenient ones. This stratified approach better models biological reality—some genes are more likely to harbor pathogenic variation and require greater scrutiny.

This method differs from both standard ACMG guidelines and previous tools. ACMG does not formally stratify frequency-based criteria by gene constraint, and most existing tools rely on fixed allele frequency cutoffs without incorporating LOEUF [28]. By dynamically adjusting cutoffs based on gene constraint, BIAS-2015 improves classification accuracy for both highly constrained and variation-tolerant genes. All cutoff values were derived from the recommended ACMG cutoffs and the updated GNOMAD LOEUF paper. These have been parameterized and exposed to the user through a constants file, where they can review and modify these values as necessary.

### ACMG Codes

BIAS-2015 evaluates 19 of the 28 ACMG codes. The remaining 9 codes can be provided through the manual adjustments step. Figure 2 shows the implemented pathogenic and benign codes and the codes that can provided through the manual step.

**Fig. 2.**
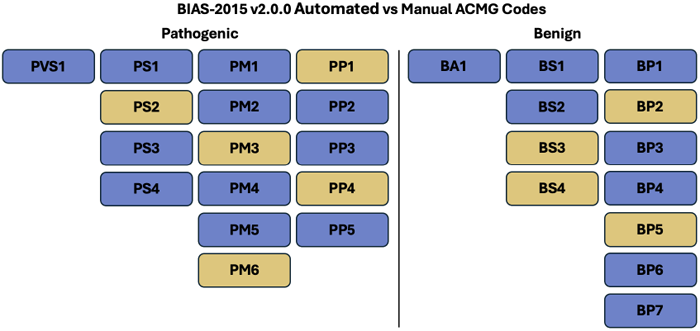
Automated vs Manual ACMG Codes. The codes in blue are automated by BIAS-2015 v2.0.0. The codes in yellow can be provided in the manual adjustment step.

BIAS-2015 does not evaluate pathogenic codes PS2, PM3, PM6, PP1, PP4 and benign codes BS3, BS4, BP2, BP5 in an automated format. These codes require patient information (EHR, parental linkage, etc) that currently is not accessible to BIAS-2015 at run time. Users can provide information for each of these classifiers during manual revisions using the external call file.

### Pathogenic Codes

#### PVS1

BIAS-2015 evaluates PVS1 by determining if a variant causes loss of function (LoF) in a gene where LoF is a known mechanism of disease. Nirvana annotates variants with their expected consequence, and for PVS1, the following consequences are considered LoF: ‘frameshift variant’, ‘stop gained’, ‘start lost’, ‘splice donor variant’, and ‘splice acceptor variant’.

If a variant is predicted to cause LoF, it is checked across all applicable transcripts [2]. If more than one transcript exists for the variant, the LoF consequence must be observed in more than one transcript for the variant to be retained. Variants in the extreme 3’ end of the final coding exon, as defined by RefSeq, are excluded [13].

Variants that meet these criteria are flagged as PVS1. BIAS-2015 then assigns a PVS1 weight based on gene constraint metrics and splicing impact predictions. If the gene has a gnomAD LOEUF is greater than 1, the variant is weighted as strong or below, as there is insufficient evidence to support a very strong classification. Splice variants with an ABSplice score indicating a low probability of pathogenicity are also weighted strong or below [11].

#### PS1 & PM5

BIAS-2015 establishes a reference dataset of pathogenic amino acid changes during preprocessing by extracting all ClinVar variants annotated as pathogenic or likely pathogenic with a review status of at least supporting strength. These variants are annotated using Nirvana to extract the gene, amino acid change, review status, and ClinVar ID, compiling a reference list of pathogenic amino acid substitutions for each gene.

When evaluating a variant, BIAS-2015 checks all P/LP ClinVar variants identified for the gene during preprocessing. If the submitted variant is identical to a reference ClinVar variant, it is skipped. Exact matches for ClinVar variants are covered under the PP5 code, so excluding them from PS1 and PM5 classification prevents using a ClinVar variant to support itself. Otherwise, if the nucleotide change differs but results in the same amino acid substitution as a known pathogenic variant, PS1 is applied. If the nucleotide change results in a different amino acid substitution at a site where a known pathogenic variant exists, PM5 is applied[31].

The review status of the reference variant determines classification strength. Variants with a supporting strength review status are assigned lower strength, while those with higher confidence, such as expert panel-reviewed variants, retain full weight.

#### PS3

This classifier looks to bring in variant classification data identified in functional studies, which requires reviewing relevant publications [29]. While traditionally a manual step, the recent emergence of artificial intelligence in bioinformatics, specifically in using natural language processing (NLP) algorithms to mine genomic data from publications, has opened access for algorithmic implementations of PS3. For the implementation of this algorithm, the authors used the UCSC AVADA (Automatic Variant Evidence Database) data track: an open-source, academic collaboration that used NLP to identify pathogenic variants from published literature [10]. We extract variants in two formats: protein and genomic. For protein variants we track the gene, the amino acid change, and the pumbedId. For genmoic variants, we track the chromosome, position, reference base, alternate base and PubMed Id.

When evaluating a variant, BIAS-2015 first searches for a direct genomic match. If no genomic match is found, it then searches for a match at the gene:protein level. If neither search yields relevant publications, PS3 is not applied. If supporting literature information is found for the variant, BIAS-2015 determines the number of unique publications referencing it. PS3 is assigned with a weight based on the number of unique publications. The PubMed IDs evaluated in PS3 are included in the output to provide traceability to the original studies.

It is possible to integrate additional data sources into this classifier, such as Genomenon Mastermind[20] or laboratory specific databases. This can be done through the pre-processing step, please reach out to the authors for advice on customization.

#### PS4

The final strong pathogenic classifier looks at genome-wide association studies to identify variants with an odds ratio (OR) indicative of genotype-phenotype relationships. We use the UCSC gwasCatalog.txt file to generate a mapping table of chromosomes to position, and position to observed GWAS score [17]. To evaluate a variant, we check if the chromosome and position are in the mapping table and extract the value if it’s present. We weight the score from supporting to strong depending on the OR and p-values, where a higher OR and lower p-value give the variant a higher score.

When GWAS data is unavailable, TOPMed, an aggregated case-control dataset, is used as a proxy for assessing allele prevalence [15]. We apply PS4 Supporting when a variant has fewer than 5 alternate alleles or between 3 and 9 homozygotes in TOPMed, considering this limited presence as suggestive of pathogenicity. This approach leverages observed allele counts in a large case-control cohort to provide supporting evidence under PS4 criteria.

#### PM1

BIAS-2015 evaluates mutational hotspots and well-established functional domains to determine whether a variant meets PM1 criteria. This classifier is implemented using ClinVar variant data and UniProt domain annotations, processed during pre-processing to identify domains enriched for pathogenic variants[5].

To define mutational hotspot domains, BIAS-2015 first builds a mapping table linking UniProt domains to ClinVar variants with a positive review status. For each domain, the algorithm calculates the percentage of variants that are pathogenic and the percentage that are benign. A domain is excluded if it contains fewer than five variants, if more than half (*>*50%) of the variants are uncertain significance, if fewer than 50% of the variants are pathogenic, or if more than 25% of the variants are benign. We exclude domains that have predominantly uncertain variants as the nature of the domain cannot be reliably ascertained. Domains that meet these criteria are saved as a required input file for BIAS-2015.

To evaluate a variant, BIAS-2015 checks whether the variant overlaps any of the identified mutational hotspot domains. If the variant is within one of these domains, it is assigned a default score of moderate. If the domain contains fewer than 75% pathogenic variants and more than 15% benign variants, the score is lowered to supporting. If the domain contains more than 90% pathogenic variants and fewer than 10% benign variants, the score is raised to strong.

#### PM2

BIAS-2015 evaluates PM2 by assessing population database allele frequencies to determine if a variant is too common to be pathogenic. This classifier relies on gnomAD and 1000 Genomes allele frequency data, extracted via Nirvana and evaluated against LOEUF specific cutoffs [4].

BIAS-2015 applies two allele frequency cutoffs for PM2: a standard cutoff (more stringent) and a supporting cutoff (less stringent). A variant is assigned supporting if its AF falls between the standard and supporting cutoffs in either gnomAD or 1000 Genomes. A variant is assigned moderate if its AF is below the standard cutoff or is completely absent from both databases.

#### PM4

BIAS-2015 evaluates in-frame insertions and deletions, which, while not as detrimental as null variants, can still indicate pathogenicity. However, in-frame indels occurring within repeat regions are less likely to be pathogenic and are excluded. Repeat regions are identified from the UCSC RepeatMasker track, intersected with UCSC consensus coding regions, and stored as a data file that is loaded at runtime.[35, 13, 1]

Variants are considered if they have one of the following Nirvana consequences: ‘stop lost’, ‘inframe insertion’, or ‘inframe deletion’. If a variant meets this criterion and is not located in a repeat region, it is assigned a base score of one. The final score is weighted based on the length of the indel: indels shorter than four amino acids are assigned a supporting strength, indels between four and ten amino acids are assigned a moderate strength, and indels longer than ten amino acids are assigned a strong strength.

#### PP2

BIAS-2015 evaulates PP2 by determining whether a missense variant is in a gene where missense variants are often associated with pathogenicity. To identify such genes, we used ClinVar data with a filtering approach similar to PM1. We created a mapping table between genes and their associated variants, then applied several exclusion criteria. If a gene had gnomAD missense observed/expected (OE) constraint data available, we excluded it if the OE value was greater than 0.4, indicating the gene is not missense constrained [14]. For genes without gnomAD OE data, we required at least one missense variant and at least one pathogenic missense variant. Genes were discarded if more than 50% of their variants were uncertain significance, if the total number of variants (pathogenic, benign, and uncertain) was fewer than five, if benign missense variants made up more than 25% of all missense variants, or if pathogenic missense variants made up less than 50% of all missense variants. The remaining genes were saved as a text file and loaded at runtime. To evaluate a variant for PP2, two conditions must be met: first, the Nirvana-assigned gene name must be in the precomputed list of genes where missense variants are the predominant pathogenic mechanism; second, the Nirvana-assigned consequence must contain the term ‘missense’. If both conditions are met, the variant is assigned a supporting score. If the OE ratio is below 10% the variant is assigned a moderate score.

#### PP3

BIAS-2015 assigns PP3 when multiple computational algorithms independently support a pathogenic classification. This classifier incorporates conservation scores (PhyloP, GERP), missense pathogenicity predictors (REVEL, DANN), and splicing impact assessments (ABSplice) [16, 25, 7, 8, 24]. Cutoff values for each tool were taken from published ClinGen recommendations [32]. If a tool’s score exceeds its ClinGen defined pathogenic cutoff, it contributes to the PP3 classification at a strength level ranging from Supporting to Very Strong, depending on how confidently it predicts a pathogenic effect. However, PP3 is immediately disqualified if any tool returns a score below the strong benign threshold.

Once all tools are evaluated, BIAS-2015 applies two post-processing rules to finalize PP3 classification. First, if only a single tool supports a pathogenic effect at the Supporting level, PP3 is not applied, ensuring that weak evidence does not lead to a pathogenic classification. Second, if multiple tools return the same strength level (e.g., two tools both indicate Moderate pathogenic evidence), the classification is upweighted by one level, as agreement between independent algorithms increases confidence. The final PP3 classification, including the tools contributing to the decision, is included in the output for transparency.

#### PP5

ACMG recommends including classification data from multiple sources and using it as the basis for the PP5 classifier [18]. This is the only pathogenic classifier that makes use of the stand-alone weighting, as we classify variants with stand-alone PP5 evidence if they have ClinVar review status ‘Practice Guidelines’ and ‘Reviewed by Expert Panel’. This ensures that variants which have already been curated and reviewed by the bioinformatics community are not falsely identified.

For remaining ClinVar variants, we reason that the evidence for their classification is not enough on it own to over rule other lines of evidence. In these situations, we provide a PP5 score consistent with our other classifiers: weighing based on evidence. Variants with a review status of ‘criteria provided, conflicting interpretations’ are assigned a score of supporting, variants with a review status of ‘criteria provided, single submitter’ are assigned a score of moderate, and for variants with a review status of ‘criteria provided, multiple submitters, no conflicts’ are assigned a score of strong.

### Benign Codes

#### BA1 & BS1

BIAS-2015 evaluates BA1 and BS1 by assessing allele frequencies in population databases to determine whether a variant is too common to be pathogenic. Both classifiers rely on gnomAD and 1000 Genomes allele frequency data, extracted via Nirvana and evaluated against LOEUF-specific cutoffs.

BA1 is assigned if a variant’s allele frequency exceeds the LOEUF-adjusted threshold in either gnomAD or 1000 Genomes, as such variants are unlikely to be pathogenic given their population frequency.

BIAS-2015 applies two allele frequency cutoffs for BS1, a standard cutoff (more stringent) and a supporting cutoff (less stringent). A variant is assigned supporting if its AF falls between the standard and supporting cutoffs in either gnomAD or 1000 Genomes. A variant is assigned moderate if its AF is below the standard cutoff or is completely absent from both databases.

#### BS2

BIAS-2015 evaluates BS2 by assessing the presence of a variant in healthy individuals using gnomAD and 1000 Genomes data, extracted via Nirvana and evaluated against LOEUF-specific cutoffs. The classification considers inheritance mode and applies distinct thresholds for autosomal recessive, autosomal dominant, and X-linked conditions identified by ClinGen Gene-Disease Validity annotations through Nirvana [26].

For autosomal recessive diseases, BS2 is assigned if the variant is homozygous in healthy individuals above the recessive homozygous threshold. If the number of homozygous observations is twice the threshold, BS2 is applied at strong strength.

For autosomal dominant diseases, BS2 is only considered in LoF-constrained genes (LOEUF below 0.5) to prevent false positive calls. A variant is excluded from BS2 if it exceeds the BS2 allele frequency threshold or if its allele count in gnomAD controls, gnomAD all populations, or 1000 Genomes all populations falls below the dominant allele threshold. If the variant is observed in at least the dominant allele threshold number of healthy individuals, BS2 is applied at strong strength. For X-linked conditions, BS2 is assigned if the variant is present in healthy hemizygous males or homozygous females above the X-linked threshold. If this condition is met, BS2 is applied at strong strength.

#### BP1

BIAS-2015 evaluates BP1 by determining whether a missense variant is in a gene where truncating variants, rather than missense variants, are typically pathogenic. To identify genes where truncating variants are the dominant pathogenic mechanism, we used ClinVar data with a filtering approach similar to PM1. We created a mapping table linking genes to their associated variants, then applied several exclusion criteria. Genes are discarded if they have fewer than one pathogenic truncating variant, more than 50% of variants are uncertain significance, less than 75% of pathogenic variants in the gene are truncating, or more than 25% of truncating variants in the gene are benign. The remaining genes are saved as a text file and loaded at runtime.

To evaluate a variant for BP1, two conditions must be met: first, the Nirvana-assigned gene name must not have been excluded by the above filters, meaning truncating variants remain the dominant pathogenic mechanism in the gene; second, the variant must not be a truncating variant, meaning its Nirvana-assigned consequence must contain the term ‘missense’. If both conditions are met, the variant is assigned a supporting score.

#### BP3

BIAS-2015 evaluates BP3 to determine whether an in-frame deletion or insertion occurs in a repetitive region without a known function. Repeat regions were identified by intersecting the UCSC RepeatMasker track with the UCSC Consensus Coding Sequence (CCDS) track, ensuring that only repeat sequences within coding regions are considered [13, 1]. To qualify for BP3, a variant must be an in-frame indel, defined as an insertion or deletion where the length is divisible by three. Additionally, variants shorter than three nucleotides are excluded unless they result in a stop lost consequence.

BIAS-2015 applies a tiered classification for BP3 based on indel length. Variants shorter than 15 nucleotides (corresponding to five amino acids) within repeat regions receive BP3 Strong, while those longer than 15 nucleotides but shorter than 60 nucleotides (19 amino acids) receive BP3 Supporting. This approach aligns with ACMG guidelines and ClinGen expert panel recommendations for evaluating in-frame indels within repetitive sequences. The final BP3 classification is included in the output, along with details of the repeat region coordinates.

#### BP4

BIAS-2015 assigns BP4 when multiple computational algorithms independently support a benign classification. This classifier incorporates conservation scores (PhyloP, GERP), missense pathogenicity predictors (REVEL, DANN), and splicing impact assessments (ABSplice). Cutoff values for each tool were taken from published ClinGen recommendations [32].

If a tool’s score falls below its ClinGen defined benign cutoff, it contributes to the BP4 classification at a strength level ranging from Supporting to Very Strong, depending on how confidently it predicts a benign effect. However, BP4 is immediately disqualified if any tool returns a score above the strong pathogenic threshold.

Once all tools are evaluated, BIAS-2015 applies two post-processing rules to finalize BP4 classification. First, if only a single tool supports a benign effect at the Supporting level, BP4 is not applied, ensuring that weak evidence does not lead to a benign classification. Second, if multiple tools return the same strength level (e.g., two tools both indicate Moderate benign evidence), the classification is upweighted by one level, as agreement between independent algorithms increases confidence. The final BP4 classification, including the tools contributing to the decision, is included in the output for transparency.

#### BP6

BP6 rules out benign variants using clinical data from ClinVar. We use the same implementation as PP5, however we check for benign classification in ClinVar instead of pathogenic.

#### BP7

BIAS-2015 applies BP7 to synonymous, intronic, or intergenic variants when computational evidence indicates that they do not affect splicing and are located in non-conserved regions. To determine BP7 eligibility, BIAS-2015 evaluates PhyloP conservation scores and ABSplice splicing predictions. Thresholds for these tools were taken from published ClinGen recommendations and the ABSplice authors’ recommended cutoffs [32].

A variant qualifies for BP7 if its PhyloP score indicates low conservation (below 7.367) and its ABSplice score is below 0.05, indicating no significant splicing impact. If the ABSplice score is below 0.01 and the PhyloP score is exceptionally low (below 0.21), BP7 is applied at Strong strength. Variants with synonymous consequences are prioritized for BP7, but intronic and intergenic variants may also qualify if they meet the same criteria. If a variant meets BP7 criteria, the classification is recorded alongside the supporting PhyloP and ABSplice scores for transparency.

### Combining Criteria

BIAS-2015 uses the ACMG combining criteria as defined with one modification. We introduce the concept of pathogenic stand-alone weighting, as a parallel to benign stand-alone weighting. BIAS-2015 only applies this weighting in PP5 to variants found in ClinVar with an ‘Expert Panel’ or ‘Practice Guidelines’ review status.

The base algorithm suggests flagging variants as uncertain when there is conflicting evidence, however they did not provide clear guidance to execute this within the combining criteria framework. Also missing was a clear order to evaluate variants, as a variant could meet both pathogenic and benign thresholds, and in this case, the order of evaluations would determine its classification.

To address these concerns, we evaluate our variants in the following order. First, we check for strong conflicting signals using a high tolerance threshold. If a variant has evidence supporting both pathogenic and benign classifications above this threshold, it is flagged as uncertain. This ensures that variants with highly contradictory evidence are not prematurely assigned a pathogenic or benign classification. Next, we evaluate whether the variant meets the criteria for a strong pathogenic classification, followed by a strong benign classification. If the variant does not meet either criterion, we conduct a second check for conflicting signals, this time using a lower threshold to identify weaker conflicting evidence. If conflicting evidence is still present, the variant is again flagged as uncertain. This second conflict check further ensures that variants with mixed but weaker evidence are not misclassified as likely pathogenic or likely benign. Finally, we assess whether the variant qualifies as likely pathogenic or likely benign. Any remaining variants that do not fit these categories are labeled as uncertain.

## Results

### Creating eRepo Derived Validation Data

ClinGen’s Evidence Repository (eRepo) serves as a pivotal resource for the clinical interpretation of genetic variants, providing expert-curated variant classifications that follow ACMG best practices. Each variant in eRepo is assessed using the ClinGen Variant Curation Interface (VCI), where expert panels apply ACMG criteria at the individual classifier level based on population data, functional evidence, computational predictions, segregation analysis, and clinical observations. These curated classifications ensure consistency, transparency, and adherence to ACMG/AMP guidelines. Importantly, eRepo contains explicitly assigned ACMG codes for each variant, along with code weights, making it an ideal reference dataset for benchmarking automated classification tools.

To evaluate the performance of BIAS-2015, we extracted 8,703 variants from eRepo and converted them into VCF format, while maintaining a linkage to their original eRepo annotation data. The final dataset, spaning 126 genes, consisted of 7,592 single nucleotide polymorphisms (SNPs), 727 deletions, 337 insertion, 49 deletion-insertion events (del-ins) and 2 MNP, ensuring a diverse representation of variant types for validation.

### Performance Analysis of eRepo Data

This dataset was then analyzed using BIAS-2015 and InterVar. We assessed the ability of each algorithm to correctly classify pathogenic, benign, and uncertain variants using eRepo as the ground truth. By comparing both algorithms to the eRepo dataset, we enable a direct comparison of sensitivity and specificity across pathogenic (P/LP), benign (B/LB), and uncertain significance (VUS) categories. The sensitivity, specificity, precision and F1 values for each code are presented in Table 1.

**Table 1.**
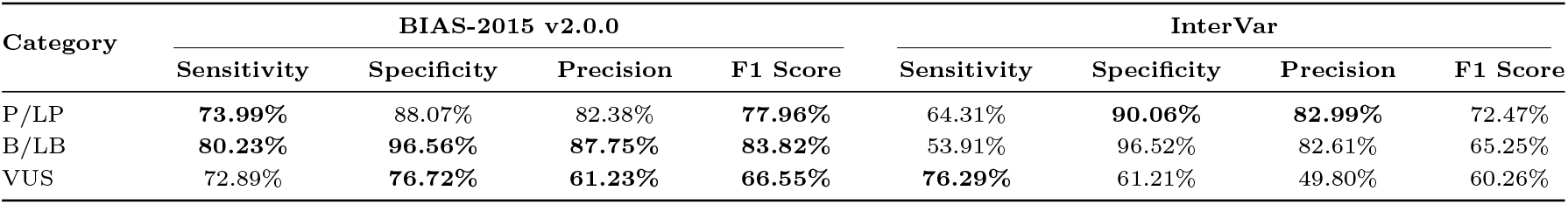
Performance Metrics of BIAS-2015 and InterVar. This table presents the sensitivity, specificity, precision, and F1 score percentages for three categories: Pathogenic/Likely Pathogenic (P/LP), Benign/Likely Benign (B/LB), and Variants of Uncertain Significance (VUS).

BIAS-2015 outperformed InterVar in overall pathogenic and benign sensitivity while maintaining comparable specificity. BIAS-2015 achieved a pathogenic sensitivity of 73.99% compared to 64.31% for InterVar, indicating improved detection of pathogenic variants. Similarly, BIAS-2015 demonstrated a benign sensitivity of 80.23%, significantly higher than InterVar’s 53.91%, while both algorithms maintained high benign specificity (96.56% vs. 96.52%). For variants of uncertain significance (VUS), InterVar showed slightly higher sensitivity (76.29% vs. 72.89%), but BIAS-2015 provided better specificity (76.72% vs. 61.21%), reducing misclassification of uncertain variants.

### ACMG Code Analysis to eRepo Data

Both BIAS-2015 and eRepo use the same ACMG classifier codes and provide written justifications for each assignment, allowing for a detailed comparison of classifier concordance. To evaluate this, we examined the number of times each classifier was assigned when processing the 8,703-variant eRepo VCF dataset and compared these counts across BIAS-2015, InterVar, and eRepo. The occurrence count for each code from each data source (eRepo, BIAS-2015 and Intervar) are shown in Figure 3.

**Fig. 3.**
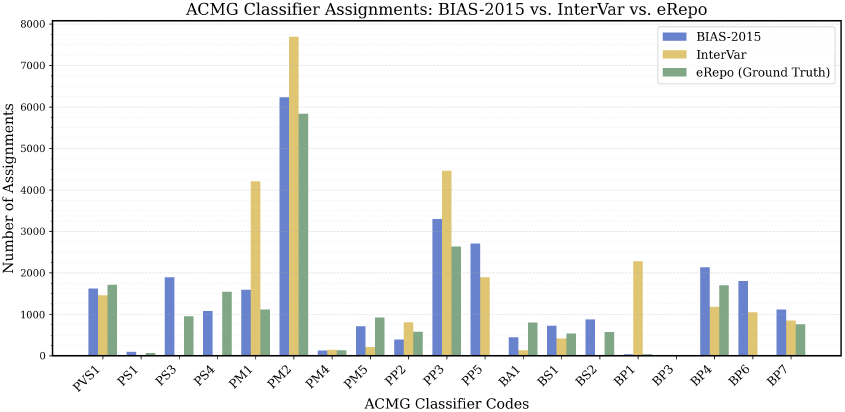
Counts of ACMG Codes by BIAS-2015, Intervar and eRepo. This bar chart compares the counts of various ACMG criteria codes between BIAS-2015 (blue), Intervar (yellow) and eRepo (green). The x-axis lists the codes, while the y-axis indicates the counts. This shows how frequently experts assign each code, and how often each algorithm assigns the same codes.

BIAS-2015 and Intervar both automate 18 ACMG codes, with BIAS-2015 additionally automating PS3. The eRepo dataset used 26 ACMG classifier codes in the classification of these 8,703 variants. The two codes excluded were BP6 and PP5, which are assigned when a variant is found in a reliable database. Since these variants were manually curated for eRepo, ClinGen reviewers did not assign these codes.

When evaluating the algorithms on the eRepo dataset, BIAS-2015 applied 19 classifiers, while InterVar assigned 16, indicating broader classifier coverage by BIAS-2015. Both BIAS-2015 and InterVar did not assign PS2, PM3, PM6, PP1, PP4, BS3, BS4, BP2, and BP5 as neither algorithm implements them. Additionally, InterVar does not assign BS2, PS3, and PS4. We note that Intervar does not evaluate PS3, however it does evaluate BS2 and PS4, and no instances of these codes were assigned in the eRepo dataset. Additionally Intervar over-flags two ACMG codes, PM1 and BP1, significantly compared to the eRepo dataset. These false positives could lead to variant missclassification.

The distribution of ACMG codes within eRepo is highly uneven, reflecting underlying biological and computational constraints rather than expert curation tendencies. Frequently applied classifiers, such as PVS1, PM2, and PP3, correspond to well-documented mechanisms of pathogenicity—loss-of-function variants in constrained genes, absence in population databases, and computational pathogenicity predictions—making them more commonly observed in variant interpretation.

Conversely, classifiers like BP3, BS4, and PS2 are rarely used, not because they are less informative, but because the necessary data to apply them is inherently limited. BP3 requires a variant to be an in-frame indel within a repetitive region, a rare genomic event. BS4 depends on large, well-documented pedigree studies, which are uncommon outside of specialized genetic cohorts. PS2, which assesses de novo status, is constrained by the availability of trio sequencing data.

For the 19 classifiers assigned by BIAS-2015, we calculated sensitivity, specificity, concordance and F1 values, all provided as supplementary data. In Figure 4 we plot the F1 scores per ACMG code of Bias and Intervar.

**Fig. 4.**
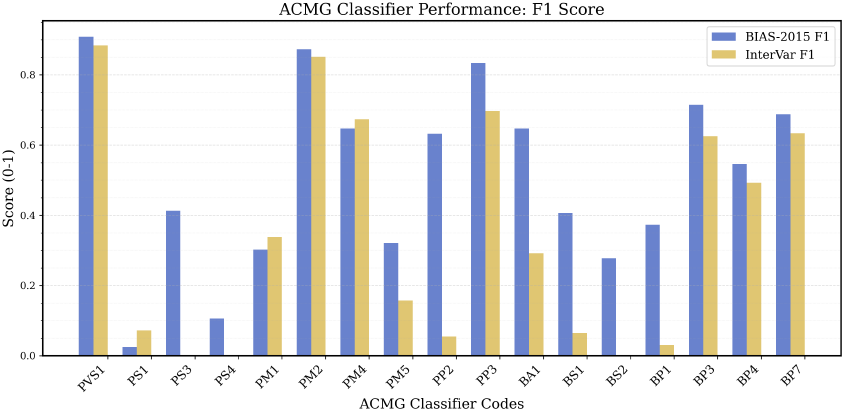
F1 of ACMG Codes from BIAS-2015 and Intervar. This bar chart compares the F1 of the 19 ACMG codes automated by BIAS-2015 (blue), Intervar (yellow). The x-axis lists the codes, while the y-axis indicates the F1 value.

BIAS-2015 demonstrates consistently higher F1 scores compared to InterVar across most ACMG classifiers, reflecting improved precision and recall. Notably, BIAS-2015 shows moderate to strong improvements over InterVar for several lower-performing classifiers, including PM5, PP2, BS2, and BP1, where InterVar struggled with high false positives and missed classifications.

Despite these improvements, both algorithms continue to show large disparities in F1 scores across different ACMG codes. High-performing classifiers such as PVS1, PM2, PM4, PP3, BP3, and BP7 consistently achieve F1 values above 0.5, while low-performing classifiers, including PS1, PS4, BS1, and BP1, remain below 0.15 for both tools. Among these, PS4 and BS1 stand out as the most critical targets for refinement due to their frequent use in eRepo classifications and their direct impact on variant interpretation. Improving PS4 requires better integration of case-control and GWAS data, while enhancing BS1 could be achieved by refining allele frequency thresholds based on gene constraint. Addressing these challenges would significantly improve overall classification accuracy.

While PP5 is derived from ClinVar and has inherent circularity concerns, it is not universally applied to all eRepo variants in BIAS-2015. Instead, it is assigned based on ClinVar’s review status and weighted accordingly. Notably, other automated ACMG classifiers such as InterVar also incorporate ClinVar data, demonstrating that the inclusion of PP5 must be evaluated in the broader context of classifier performance rather than in isolation.

### Speed Analysis of ClinVar Data

To evaluate the speed of BIAS-2015, we tested the complete annotation pipelines for InterVar and BIAS-2015 on the full ClinVar VCF file from January 30, 2025, containing 3,040,405 variants. All analysis was performed on an AWS LightSail cloud instance with CPU: Intel Xeon Platinum 8175M @ 2.50GHz (4 cores, 8 threads) and 30 GiB of RAM. Nirvana took 18 minutes and 24 seconds to annotate the variants, and BIAS-2015 classified them in 19 minutes and 47 seconds, achieving an average processing speed of 1,327 variants per second. Conversely, ANNOVAR took 1 hours, 20 minutes and 19 seconds to annotate the variants, and InterVar required an additional 6 hours, 4 minutes and 39 seconds to classify them, for an average of 113 variants per second. This represents an 11.5x increase in speed with BIAS-2015 compared to InterVar, demonstrating its efficiency for high-throughput variant classification without sacrificing accuracy.

### Interactive Web Application for Variant Curation

To further enhance the usability of BIAS-2015, we developed a React-based web application designed to facilitate genomic variant classification using ACMG criteria. This interactive platform, shown in Figure 5, provides a comprehensive interface for visualizing, managing, and curating variant data, ensuring efficient documentation and modification of classifications with supporting evidence. Built with modern web technologies (React, TypeScript, and Tailwind CSS), the application enables tabular variant display, interactive ACMG criteria management, and seamless data import/export.

**Fig. 5.**
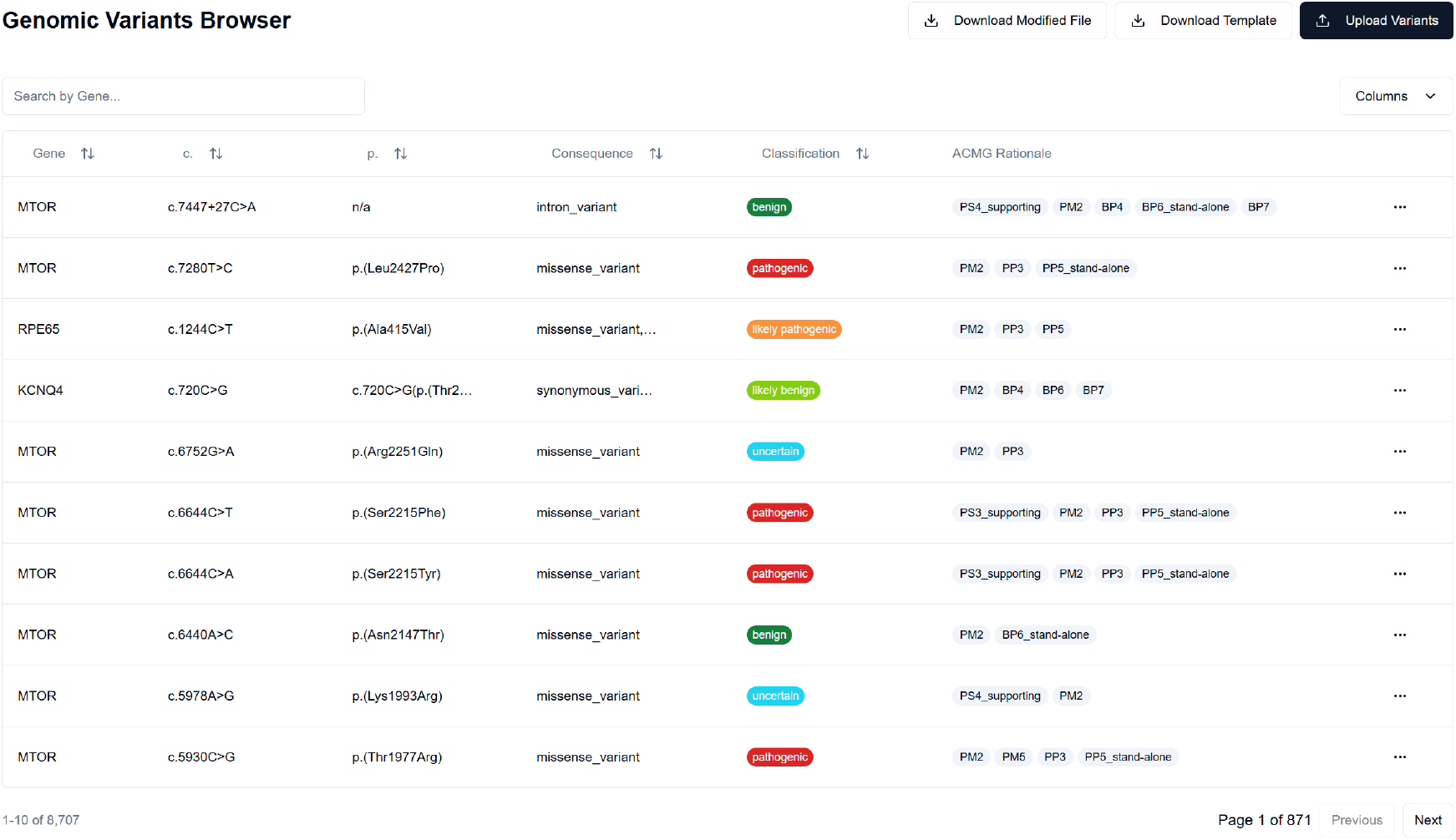
BIAS-2015 Genomic Variants Browser. This screenshot displays a subset of MYH7 variants classified by BIAS-2015, showing the ACMG classification, supporting criteria, and available actions for manual review. The interactive UI enables researchers and geneticists to efficiently manage variant curation.

The UI was designed to complement BIAS-2015’s high-throughput capabilities, allowing users to review and refine classifications efficiently in both research and clinical settings. By integrating an interactive ACMG criteria framework, the application supports real-time classification adjustments, making it particularly valuable for variant curators, geneticists, and researchers working with large genomic datasets. This interface ensures transparency in variant interpretation while maintaining compliance with ACMG/AMP guidelines, bridging the gap between automated classification and expert review.

## Discussion

### Manual Adjustment

While BIAS-2015 performs automated variant classification, the authors emphasize that manual review remains an essential component of accurate interpretation. Certain ACMG classifiers that BIAS-2015 does not evaluate must be provided by the user, and in cases where additional clinical or research insights are available, users may wish to adjust classifier weights. Previously, this process required modifying an external call file, but with the BIAS-2015 web interface, users can now directly modify classifications within the UI.

The interactive interface allows users to upload variant files, manually edit ACMG classifier assignments, adjust weights, and save the modified classifications for downstream analysis.

This replaces the previous workflow, where users had to modify the external call file manually in JSON or TSV format before rerunning the algorithm. The UI simplifies this process by providing a structured, intuitive way to edit individual variant classifications in real-time.

Figure 6 displays an example of the ACMG Classification Details panel within the BIAS-2015 UI. Users can review the automatically assigned ACMG criteria, modify their strengths, and input additional supporting evidence. Changes can be saved and re-exported, ensuring that variant curation remains flexible and transparent while maintaining traceability.

**Fig. 6.**
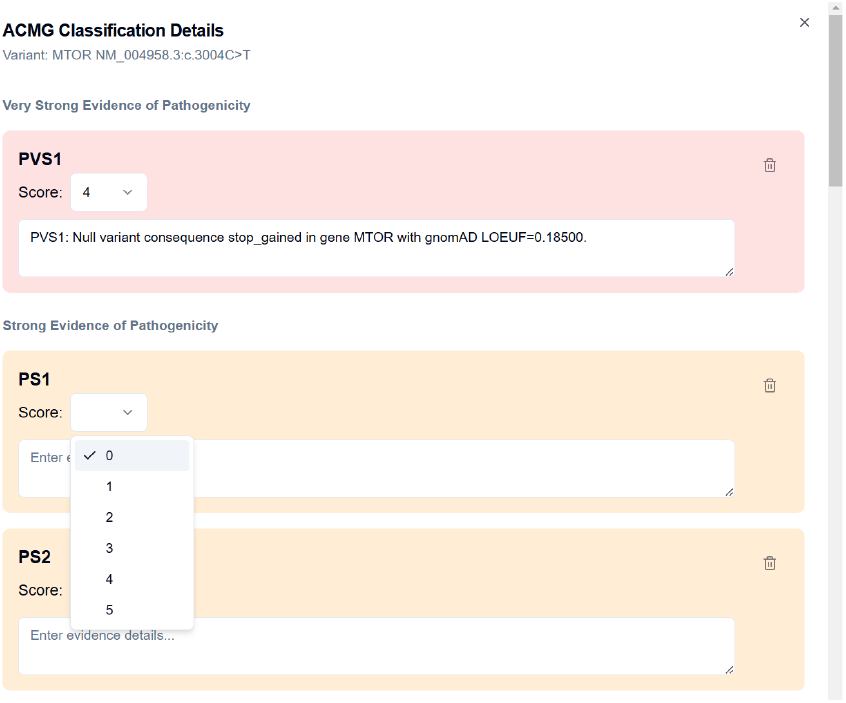
BIAS-2015 ACMG Classification Modification Interface. This screenshot shows the manual classification adjustment panel, where users can review, modify, and document ACMG classifier assignments. The interface provides structured editing for both automated and manually assigned classifications, supporting expert curation and review.

### Limitations and Next Steps

While BIAS-2015 demonstrates strong performance in automating ACMG classification, further refinements are necessary to improve challenging classifiers and optimize classification accuracy. Some ACMG criteria remain inherently difficult to automate due to their reliance on limited or complex datasets. For example, PS4 (case-control and GWAS data) and BS1 (benign allele frequency thresholds) currently have lower F1 scores and require additional refinements to improve precision. Enhancing PS4 will require deeper integration of large-scale GWAS datasets and population-based allele frequency modeling, while BS1 can be improved by refining allele frequency cutoffs in gene-specific contexts.

Another area of ongoing improvement is the stratification of LOEUF-based allele frequency thresholds. While BIAS-2015 dynamically adjusts frequency cutoffs based on gene constraint, further refinement is possible by incorporating additional gene-level metrics, such as essentiality scores and tissue-specific expression constraints. Future versions of BIAS-2015 will aim to refine population frequency-based classifiers (e.g., PM2, BS1, and BA1) by implementing a more granular LOEUF-based stratification model.

To ensure transparency and adaptability, all cutoff values have been parameterized into a constants file, which users may review and modify as needed. Where possible, these cutoffs were derived from published sources; otherwise, they were determined based on experimental evidence. Additionally, validation code is available to assess sensitivity, specificity, and F1 scores, along with scripts to generate validation datasets. This allows users to fully vet any modifications they implement.

Lastly, BIAS-2015’s dynamic weighting system provides an important advancement in ACMG classification, but additional refinements are needed to better calibrate classifier strengths. Future efforts will focus on further optimizing weight adjustments based on empirical validation against expert-curated datasets. Additionally, integrating machine learning models to refine classifier weights based on large variant datasets may further enhance classification performance.

By addressing these challenges, BIAS-2015 will continue to evolve, providing increasingly accurate and interpretable variant classifications while maintaining alignment with ACMG/AMP guidelines.

## Supporting information

complete_per_code_performance_analysis

## Data Availability

The algorithm code, pre-processing scripts, validation tools, image generation code, and manuscript creation resources are openly available on GitHub to ensure transparency and accessibility for the broader scientific community.
Algorithm Code and Resources: https://github.com/bitscopic/BIAS-2015
GUI Code: https://github.com/bitscopic/BIAS-2015-UI
Precomputed required files for BIAS-2015 v2.0.0 are available via AWS S3:
HG19 Data Files: s3://bias-2015/bias_v2.0.0_hg19_data_files.zip
HG38 Data Files: s3://bias-2015/bias_v2.0.0_hg38_data_files.zip
All supplementary files, including the validation VCF and intermediate validation files used in the manuscript, are available at:
s3://bias-2015/bias_v2.0.0_supplementary_files.zip
Data Availability Statement:
All data used in this study are openly available through the sources listed above. No restrictions apply to data access.

https://github.com/bitscopic/BIAS-2015-ui

## Declaration of Interests

The authors declare the following conflict of interest: The BIAS-2015 algorithm described in this paper is intended for use in a commercial product by Bitscopic. This potential financial interest does not alter the authors’ adherence to community best practices and scientific standards in developing and validating the algorithm. The development of BIAS-2015 was conducted independently by Bitscopic, and no external funding or influence was received from commercial entities outside of this potential interest.

## Author Contributions

- CE: Conceptualization, Methodology, Software, Validation, Writing, Review & Editing.
- RB: GUI, Writing, Review & Editing.
- BN: Software, Validation & Review.
- JM: Software, Review & Editing.
- VB: Writing & Review.

## Data and Code Availability

The algorithm code, pre-processing scripts, validation tools, image generation code, and manuscript creation resources are openly available on GitHub, ensuring transparency and accessibility for the broader scientific community.

**Algorithm Code and Resources**

https://github.com/bitscopic/BIAS-2015

**GUI Code**

https://github.com/bitscopic/BIAS-2015-UI

**Precomputed Required Files for BIAS-2015 v2.0.0**

`s3://bias-2015/bias v2.0.0 hg19 data files.zip s3://bias-2015/bias v2.0.0 hg38 data files.zip`

**Supplementary Files:** All supplementary files, including the validation VCF and intermediate validation files used in the manuscript, are available here:

`s3://bias-2015/bias v2.0.0 supplementary files.zip`

---

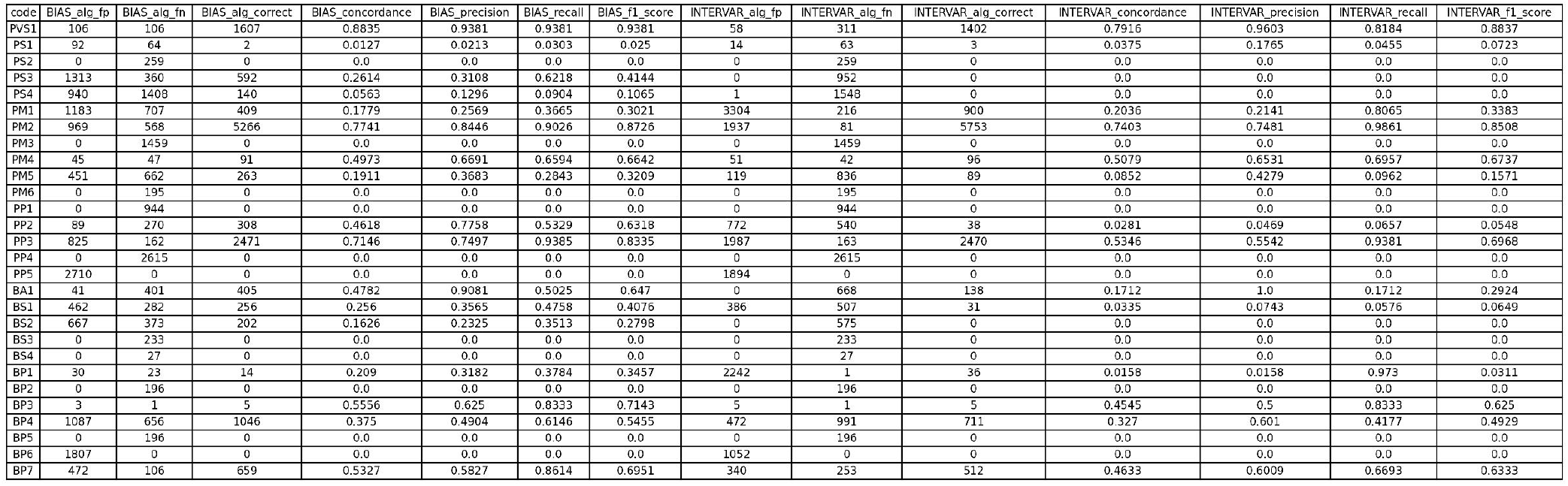

## Notes

### Author Declarations

The study used ONLY openly available human data that were originally located at: UCSC Genome Browser public datasets: RefSeq: https://hgdownload.soe.ucsc.edu/goldenPath/hg19/database/ncbiRefSeqHgmd.txt.gz RepeatMasker: https://hgdownload.soe.ucsc.edu/goldenPath/hg19/database/rmsk.txt.gz CCDS: https://hgdownload.soe.ucsc.edu/goldenPath/hg19/database/ccdsGene.txt.gz GWAS Catalog: https://hgdownload.soe.ucsc.edu/goldenPath/hg19/database/gwasCatalog.txt.gz AbSplice: https://hgdownload.soe.ucsc.edu/gbdb/hg19/abSplice/AbSplice.bb AVADA: http://hgdownload.soe.ucsc.edu/gbdb/hg19/bbi/avada.bb UniProt Domains: https://hgdownload.soe.ucsc.edu/gbdb/hg19/uniprot/unipDomain.bb gnomAD Missense Constraint: https://hgdownload.soe.ucsc.edu/gbdb/hg19/gnomAD/missense/missenseConstrained.bb ClinVar dataset (NCBI): https://ftp.ncbi.nlm.nih.gov/pub/clinvar/vcf_GRCh37/clinvar.vcf.gz Illumina Nirvana (v3.18.1): https://github.com/Illumina/Nirvana/releases/tag/v3.18.1 FDA-approved eRepo database (ClinGen Evidence Repository): https://erepo.clinicalgenome.org/evrepo/ All datasets used in this study are fully public, do not require any registration, licensing, or access approval, and were openly available before the initiation of the study.

